# Catchii: empowering literature review screening in healthcare

**DOI:** 10.1101/2023.02.10.23285791

**Authors:** Andreas Halman, Alicia Oshlack

## Abstract

A systematic review is a type of literature review that aims to collect and analyse all available evidence from the literature on a particular topic. The process of screening and identifying eligible articles from the vast amounts of literature is a time-consuming task. Specialized software has been developed to aid in the screening process and save significant time and labour. However, the most suitable software tools that are available often come with a cost or only offer either a limited or a trial version for free.

In this paper, we report the release of a new software application, Catchii, which contains all the necessary features of a systematic review screener application while being completely free. It supports a user at different stages of screening, from detecting duplicates to creating the final flowchart for a publication. Catchii is designed to provide a good user experience and streamline the screening process through its clean and user-friendly interface on both computers and mobile devices, as well as features such as multi-coloured keyword highlighting, the ability to screen titles and abstracts smoothly with an unstable or even absent internet connection, and more.

Catchii is a valuable addition to the current selection of systematic review screening applications that also allows researchers without financial capabilities to access many of the features found in the best paid tools. Catchii is available at https://catchii.org

## Introduction

A systematic literature review or systematic review (SR) is a research method used to systematically evaluate the scientific literature on a specific topic. The process involves identifying relevant studies from a large pool of literature, assessing their relevance and quality, and synthesizing their findings to provide an overview of the current state of knowledge on the topic. SRs are commonly used in the healthcare field and are becoming increasingly popular, with a four-fold increase in the number of SRs published over the past decade indexed by PubMed with over 30 thousand articles in 2022 alone [1].

One of the challenges of conducting an SR is the time-consuming process of screening articles. To address this, various software tools and applications have been developed to aid reviewers during the screening process. A feature analysis of 16 different SR software applications and methods was conducted by Van der Mierden and colleagues, who determined and scored the mandatory, desirable and optional features of an SR tool in the biomedical research context [2]. The five highest-scoring tools were DistillerSR [3], EPPI-Reviewer [4], SWIFT Active Screener [5], Covidence [6] and Rayyan [7]. The authors concluded that the first four applications all support mandatory features and are preferred for screening [2]. However, those applications require a paid subscription. The highest scoring free application was Rayyan, but it does not have all the mandatory features, specifically missing the full-text screening phase [2]. Similarly, another comparison of SR tools in healthcare recommended Covidence and Rayyan as the highest-scoring tools in their feature analysis [8].

With regards to pricing, as of February 2023, the cheapest DistillerSR package costs $19.95 (USD) a month for three students (limited to one project) and access to multiple projects and additional features requires a more expensive package [3]. EPPI-Reviewer provides two packages: £10/month ($12.40, based on the current exchange rate) for one user (unlimited single-user reviews) and £35/month ($43.40) for one user (unlimited multiple-user reviews) [4]. Covidence on the other hand has yearly packages, the cheapest one costs $240 for one user and is limited to one review [6]. While Rayyan is commonly considered as a free tool, the free version is limited. For example, it has a maximum of three active reviews, a maximum of 100 decisions on mobile app, cannot revoke a reviewer and no PRISMA flow-chart generation is included. To access more features from Rayyan requires a professional package that costs $8.25/month for one user [7]. All in all, a substantial cost can be incurred by researchers as a systematic review takes around 15 months on average to complete and publish [9], and the use of two reviewers for screening is the current standard to prevent missing papers [10].

In this article, we present a new software application called Catchii, which is designed to perform efficient and fast screening of articles for a systematic review. The development of Catchii focused on creating a user-friendly experience and included all necessary features identified for an SR tool. Importantly, Catchii is offered as a free unlimited application, enabling researchers without financial resources to access the same features as paid applications, facilitating more equitable access for conducting an SR.

## Design and implementation

### Application design and selection of features

We designed Catchii to include all the features of importance in SR software defined in the literature screening tool comparison analysis [2]. This article rates the importance of features into three categories: mandatory, desirable and optional. The features are related to the technical functioning of the tool and include: support for importing references (M1), multiple users (M2), the ability to (re-)allocate references (M3), having distinct title-abstract (TiAb) and full-text phases (M4), including or excluding references (M5), resolving discrepancies (M6), exporting results (M7), having a stable release of the software (M8) and the level of customer support (M9). The desirable and optional features are not critical for a SR, but they can assist the process. Features such as support for non-Latin characters (D1), multiple different user roles (D2), randomising the order of references (D3), keyword highlighting (D4), attaching PDF (Portable Document Format) files (D5), attaching comments (D6), displaying project progress (D7), project auditing (D8), and being free to use (D9) were considered desirable, while reference labelling (O1), some form of machine learning/automation (O2) and flow-diagram creation (O3) were considered optional. An acceptable threshold was assigned for each feature (for example, a “distinct TiAb and full-text” feature has the following options: “no distinct phases”, “TiAb & full-text phase” and “user defined phases” and acceptable threshold is “TiAb & full-text phase”). In the following sections the Catchii implementation of each feature at acceptable threshold is discussed.

### Implementation of features

Catchii is a systematic literature review tool that fully supports reference importing (M1). Users can easily import references in different formats, including files created by PubMed, Web of Science, OVID (e.g. PsycINFO, MEDLINE), Scopus, a general RIS (Research Information Systems) file created by online databases or reference manager tools, as well as a CSV (Comma Separated Values) file. For each uploaded set, a dataset label can be specified for the contained references (O1). Additionally, Catchii supports articles written using various characters (D1), including Latin, Cyrillic, Chinese, Japanese, Arabic and others.

Each user can perform screening for an unlimited number of reviews, and for each review, multiple users are supported (M2). Each reviewer is blind to others’ decisions until everyone has made their decision for a particular article. Multiple roles for reviewers (D2) are also supported, including a manager of the review (with full access to the review), a reviewer (with rights to make inclusion/exclusion decisions but cannot resolve discrepancies), a reviewer with the rights to also resolve discrepancies, and a reviewer who only has rights to review and resolve discrepancies (if there are any).

In the event that a reviewer drops out, references from that user can be re-allocated (M3) to a new user. Catchii has an easy system for adding, removing, and replacing reviewers from a review. A replacement reviewer can be added to the review at any time including before other reviewers have finished the title-abstract screening.

Catchii also has a built-in duplicate detection feature which compares the title or abstract of all articles published in a particular year by using the similar_text() function in the “PHP: Hypertext Preprocessor” language to calculate similarity in percentages between two text strings. Articles that have an abstract or title that is at least 80% similar are returned to the user for review. By default, all duplicate records except one are marked and can be collectively removed with one click. In the case of a missing abstract, only titles are compared. We consider this feature as an automation feature (O2), as users do not have to manually find duplicate records but can automate the task. However, from the Van der Mierden article it is unclear which automation features are included in this category [2].

Catchii also supports distinct TiAb and full-text phases (M4). In both phases, reviewers must decide whether to include or exclude a particular article (M5). In the full-text phase, a reason for exclusion can be recorded by the reviewer. Each phase is divided into two parts: in the first part, inclusion/exclusion decisions are made, and in the second part, the decisions are confirmed and discrepancies are resolved (M6). Technically, a decision for a disagreement between reviewers can be made by the manager of the review, a reviewer with rights to solve disagreements, or an additional reviewer who only has rights to solve disagreements and does not take part in the screening process. In the screening process, each reviewer can choose the order of records: either sort them by year, title, dataset label, or list them in a randomized order (D3).

To enhance the efficiency of title and abstract assessment, Catchii has incorporated a keyword highlighting feature (D4). In contrast to most systematic review tools, Catchii supports multiple groups of keywords and allows for the assignment of distinct colours to differentiate them. Conventional systematic review tools typically utilize two sets of keywords – inclusion and exclusion, which are highlighted in green and red, respectively. This approach can limit the acquisition of information and may cause difficulties for individuals with red-green colour blindness. By utilizing keyword groups, reviewers are able to assign different colours for inclusion or exclusion keywords. For example, when the inclusion keywords are “Sun,” “UV,” “cancer,” and “humans,” Catchii can assign distinct colours for each keyword, allowing for quick visual acquisition. We believe that this improves the assessment of abstracts and ultimately accelerates the screening process. The colours used are chosen from a range of the colour spectrum and are suitable for individuals with different forms of colour blindness. Additionally, reviewers may also have the ability to collect papers for other purposes during the screening process, which may not be included in the review but are still valuable to the reviewer. By specifying a keyword group to identify these papers, a reviewer can then add them as a favourite (an additional feature of Catchii) and exclude from the screening process, but later return to the list of favourite articles.

In the full-text phase, Catchii supports the attachment of PDFs (D5) both manually and through automatic download from publicly available resources on the Internet, such as PubMed Central. Uploaded PDFs will be accessible to all reviewers. If necessary, each reviewer can add comments (D6) to the records that can be made visible to other reviewers or kept private for the writer. A detailed project progress (D7) is displayed for each project/review, separated by phases and sub-phases. The progress of each reviewer can be also viewed by other co-reviewers on the review.

Upon completion of the screening, records can be fully exported (M7) into a reference manager software or as a CSV file. PDF files can be added to the exported references and opened in a reference manager software, making it easy to perform data extraction on the user’s computer. A PRISMA flowchart can be generated (O3) as an editable document, allowing reviewers to make any necessary corrections. Catchii has a built-in logging system where each decision and update in a review is recorded. For auditing purposes, the manager of the review can download an audit log (D8) to review the actions of each reviewer.

Finally, Catchii is a free-to-use tool (D9) with a stable release (M8) that is actively supported. There is a short documentation, a forum, and a direct support option for users (M9). Additionally, Catchii has other features not previously outlined in the requirement from Van der Mierden. Specifically, Catchii supports mobile devices, free-text data extraction, the ability to bookmark articles into a favourites list, and an offline screening mode that enables screening of records even without an internet connection, such as on airplanes or in areas with an unstable connection.

## Results

### Duplicate detection

We evaluated the performance of duplicate detection of Covidence and Rayyan, tools which are one of the most popular and have been recommended [2, 8]. Both tools were compared to Catchii, in two ways. Firstly, we assessed their sensitivity by counting the number of duplicates detected. We conducted a search for records containing the term “7,8-DHF” in PubMed, Scopus, and Web of Science databases, and selected all records published between 2010 and 2020 (inclusive). A total of 390 records were downloaded (130 in PubMed, 125 in Scopus, and 135 in Web of Science) [11]. We manually reviewed the records and identified 246 duplicates in the combined dataset. Then, we imported the records into Rayyan, Covidence, and Catchii, and counted the number of duplicates detected by each tool.

Catchii missed four records out of 246 (98.4% sensitivity) - one with a different publication year and three records with a large block of text added to the abstract that resulted in a low similarity percentage, below the 80% threshold. Catchii showed higher sensitivity than Covidence, which missed eight records (94.3%), but had lower sensitivity than Rayyan, which found all duplicates (100%). However, while Rayyan did not miss any duplicates, their system requires manual resolution of duplicates, which is a time-consuming task. For example, resolving 100 records with one duplicate each would require 300 clicks (select the record, click on a link to resolve the duplicate and click to remove the chosen duplicate). In Covidence, duplicates are removed automatically after import, with the review of removed duplicates taking place afterwards. In Catchii, all duplicates can be removed with one click after reviewing them.

Secondly, we evaluated the duplicate detection capabilities of each tool. For this purpose, we downloaded six records and modified them manually to test various scenarios [11]. We tested case sensitivity by converting one record’s abstract and title to uppercase, altered the abstract (added a short copyright notice in the end), created duplicates with more than two instances of the same record, removed abstract of one record (titles were identical), removed abstract of one record and altered title (added “[review]” in the end), and the use of non-Latin characters (title and abstract both in Mandarin Chinese).

Of the three tools, only Catchii was able to accurately identify all types of duplicates (6/6). Both Rayyan and Covidence missed the duplicate that had a missing abstract and a small alteration in the title (5/6). The results of both experiments are presented in Table 1.

**Table 1.** Results of duplicate detection by Rayyan, Covidence and Catchii.

|  | **Rayyan** | **Covidence** | **Catchii** |
| --- | --- | --- | --- |
| Sensitivity | 246/246 (100%) | 238/246 (94.3%) | 242/246 (98.4%) |
| Different types of duplicates detected | 5/6 | 5/6 | 6/6 |
| Resolving duplicates | Very slow<br>(manual resolving) | Fast<br>(removed automatically) | Fast<br>(one-click removal) |

### Feature comparison

The systematic review screening tools comparison article by Van der Mierden evaluated available software based on the number of features they offer in three categories [2]. We calculated the score for Catchii by taking into account the number of features with an acceptable threshold described previously and combining it with the results of the Van der Mierden article. Catchii received a score of 20 points (21, if duplicate detection and removal is considered automation), which is the same or higher than the highest-scoring tool, DistillerSR. Compared to Catchii, DistillerSR has one fewer desirable feature but one more (or the same) optional features. As shown in Table 2, among the top-scoring SR tools, Catchii is the only one that is completely free for the user.

**Table 2.**
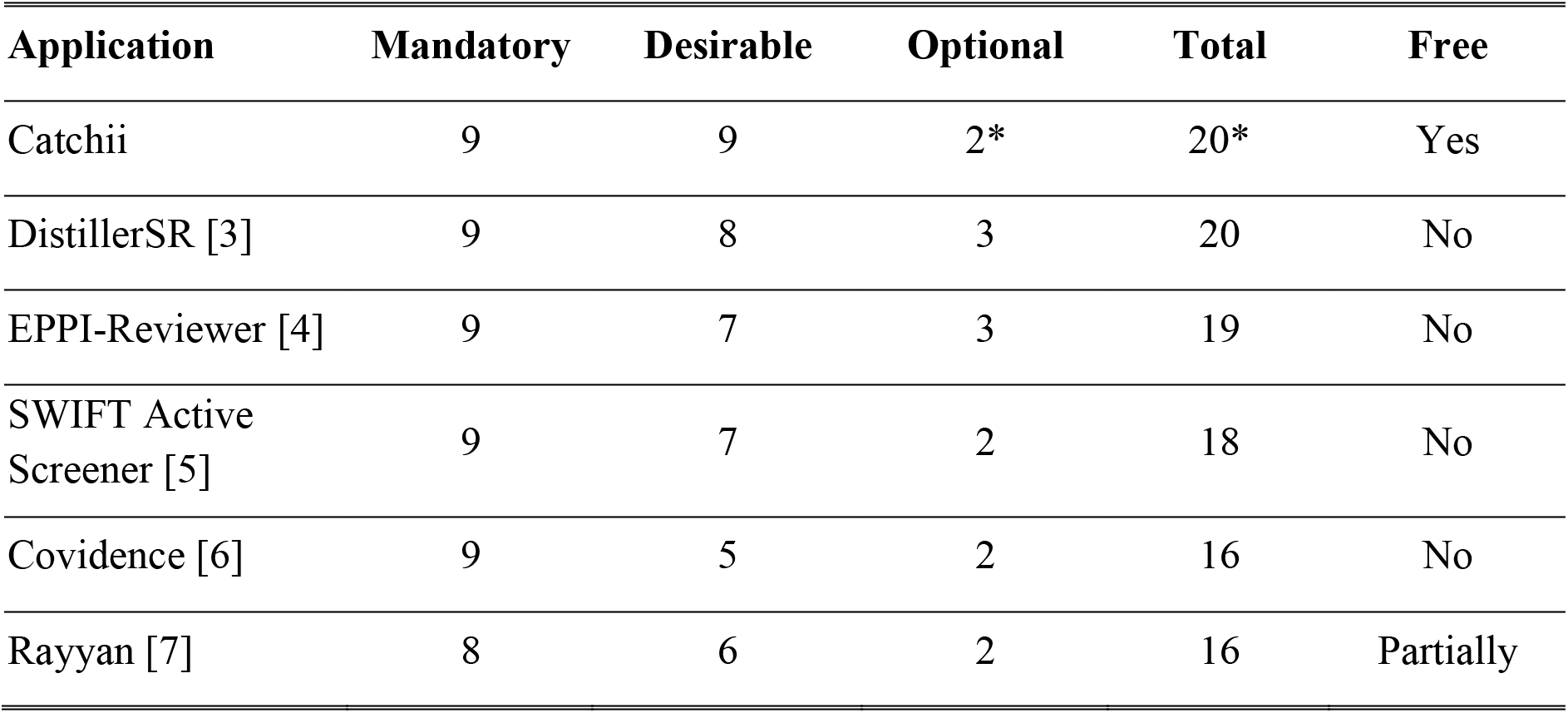
Feature comparison of SR tools. The number of features as per Van der Mierden, et al. article [2] including the addition of Catchii, and an indication of whether the software is free. In the article, the first completely free tool (CADIMA) scored 14 points in total and the next partially free tool (SysRev) scored 15 points, positioning right after Rayyan (not shown). The score in the “optional” features is either 2 or 3 (and total score of 20 or 21) depending on whether duplicate detection and removal is considered as automation, which remained unclear from the Van der Mierden article.

| **Application** | **Mandatory** | **Desirable** | **Optional** | **Total** | **Free** |
| --- | --- | --- | --- | --- | --- |
| Catchii | 9 | 9 | 2* | 20* | Yes |
| DistillerSR [3] | 9 | 8 | 3 | 20 | No |
| EPPI-Reviewer [4] | 9 | 7 | 3 | 19 | No |
| SWIFT Active Screener [5] | 9 | 7 | 2 | 18 | No |
| Covidence [6] | 9 | 5 | 2 | 16 | No |
| Rayyan [7] | 8 | 6 | 2 | 16 | Partially |

Additionally, we calculated a score for the Systematic Review Accelerator (SRA) tool [12], which was published after Van der Mierden’s article and thus was not included in the comparison. The SRA toolbox is not an all-in-one screening application, but more a set of tools (Deduplicator, Screenaton and Disputatron) that can be used individually to perform deduplication, TiAb screening and resolving disagreements between reviewers (respectively), which can be evaluated based on their features. The following features with an acceptable threshold were found for the SRA: M1, M2, M3 (yes/no; unsure), M6, M7, M8, M9, D1, D4, D9, and O2 (Deduplicator), resulting in a total score of 10 or 11, and thus it was not included in the comparison table with the top five tools.

## Discussion

Catchii is a systematic review screening application, designed with a user-focused approach and optimized for use on computers, tablets, and mobile devices. It contains all the essential features of SR screening software, as well as numerous desirable and optional ones, making it a whole solution for reviewers. We have demonstrated that Catchii offers equal or more features to the leading SR screening tools available, while also being free to use.

Catchii has some recognised limitations. Firstly, it is oriented to health sciences and therefore currently supports the main formats of records in biomedical literature and thus may not have the capacity to directly import records from some databases used in other fields. Although this issue can be overcome by using CSV files, we aim to expand our support for additional formats based on user demand. Secondly, our duplicate detection algorithm compares articles published in the same year only. This significantly speeds up the process but could result in missing some duplicates whose publication year differs between databases. However, as is rather a rare occurrence (for example, only one duplicate with a different year was present in our test dataset) it is likely faster to review a few additional undetected duplicate records for the whole dataset than wait for a much slower deduplication process to finish.

Catchii’s support and development is ongoing and we have plans to introduce more features to assist users in their systematic review process. The application can be accessed from https://catchii.org

## Data Availability

The datasets generated and analysed in the current study are available in the Zenodo repository, https://doi.org/10.5281/zenodo.7613707

## Declarations

### Competing interests

The authors declare that they have no competing interests.

### Funding

AO is funded by an NHMRC Investigator Grant GNT1196256.

### Authors’ contributions

AH conceived the project, designed and developed the software, assessed the duplicate detection systems of tools and drafted the manuscript. AO advised on different stages of the project and revised the manuscript. Both authors read and approved the final manuscript.

## List of abbreviations

CSV: Comma Separated Values
PDF: Portable Document Format
RIS: Research Information Systems
SR: Systematic Review
SRA: Systematic Review Accelerator
TiAb: Title and Abstract
USD: U.S. Dollar

## Notes

### Competing Interest Statement

The authors have declared no competing interest.

## References

1. NCBI PubMed. In: NCBI PubMed. https://pubmed.ncbi.nlm.nih.gov/. Accessed 26 Jan 2023

2. Van der Mierden S, Tsaioun K, Bleich A, Leenaars CHC (2019) Software tools for literature screening in systematic reviews in biomedical research. ALTEX 36:508–517

3. DistillerSR Inc DistillerSR. https://www.distillersr.com/. Accessed 7 Feb 2023

4. EPPI-Centre EPPI-Reviewer: systematic review software. https://eppi.ioe.ac.uk/cms/Default.aspx?alias=eppi.ioe.ac.uk/cms/er4. Accessed 7 Feb 2023

5. Sciome LLC SWIFT Active Screener. https://www.sciome.com/swift-activescreener/. Accessed 7 Feb 2023

6. Veritas Health Innovation Covidence systematic review software. https://www.covidence.org. Accessed 7 Feb 2023

7. Rayyan. https://www.rayyan.ai. Accessed 7 Feb 2023

8. Harrison H, Griffin SJ, Kuhn I, Usher-Smith JA (2020) Software tools to support title and abstract screening for systematic reviews in healthcare: an evaluation. BMC Med Res Methodol 20:7

9. Borah R, Brown AW, Capers PL, Kaiser KA (2017) Analysis of the time and workers needed to conduct systematic reviews of medical interventions using data from the PROSPERO registry. BMJ Open 7:e012545

10. Waffenschmidt S, Knelangen M, Sieben W, Bühn S, Pieper D (2019) Single screening versus conventional double screening for study selection in systematic reviews: a methodological systematic review. BMC Med Res Methodol 19:132

11. Halman A (2023) Records for determining the sensitivity and capabilities of duplicate detection systems [dataset]. https://doi.org/10.5281/zenodo.7613707

12. The Systematic Review Accelerator (SRA). https://sr-accelerator.com/. Accessed 7 Feb 2023

